# Conducting MRI trials in Alzheimer’s patients: Challenges and Guidelines

**DOI:** 10.1101/2025.07.02.25330723

**Authors:** Maria Dobrushina, Sayyeda Chandni, Linda Dietrich, Wenzel Glanz, Michaela Butryn, Dorothea Hämmerer, Lucía Penalba-Sánchez

## Abstract

Combining pharmaceutical interventions with neuroimaging in Alzheimer’s disease (AD) research presents logistical and methodological challenges. This perspective paper outlines early-phase experiences from a 7-tesla (7T) fMRI clinical trial involving individuals with mild cognitive impairment (MCI) and mild AD, highlighting recruitment hurdles and practical recommendations. From a pool of 1,000 patients, 475 had MCI or AD clinically diagnosed; following pre-selection and screening, only 6% met all inclusion criteria. Major barriers included exclusion due to comorbidities, difficulties with blood draws, miscommunication about study procedures, and unreported medical conditions discovered during MRI. Effective communication, often requiring caregiver involvement, was essential for obtaining accurate medical histories and improving adherence. Internally, clear team coordination helped manage scheduling and protocol compliance. While strict eligibility criteria enhance data quality, they significantly impede recruitment and feasibility in high-field MRI drug studies. We offer recommendations to optimize recruitment and screening, improve protocol execution, and better balance scientific rigor with real-world clinical constraints.

## 1 | Background

Alzheimer’s disease is the most prevalent cause of dementia worldwide currently affecting around 55 million individuals worldwide[1]. Although numerous clinical and MRI studies are underway to better understand this disease and delay the onset of dementia, their progress is typically slow, often taking several years to acquire high-quality, well-characterized samples[2][3]. As a result, researchers may be tempted to relax inclusion criteria to meet project timelines, potentially compromising the robustness and generalizability of study findings[4].

Key aspects of a clinical study’s early phases, such as study design, eligibility criteria, recruitment, medical screenings, and data collection, along with essential transversal skills like biological and medical data management, team coordination, and communication with participants and staff, are often overlooked and underreported. Yet, these factors significantly influence the study’s efficiency, and overall quality.

Conducting a study including a pharmaceutical drug, MRI and an older population with AD in our lab presented numerous demands. This article provides a behind the scenes look of the study. More specifically, it offers a qualitative and a descriptive statistical analysis of the “backstage” processes and challenges encountered during the initial phases of a 7T fMRI study investigating the effects of atomoxetine on the noradrenergic system. Additionally, it highlights crucial aspects that contributed to our study’s success and offers guidelines for future studies (see Supplementary Material for further details on the study).

## 2 | Methods

In this manuscript *first*, we describe the recruitment process, including pre-selection of participants via our internal memory clinic database at the German Center for Neurodegenerative Diseases (DZNE) - University hospital Magdeburg clinic for neurology; and telephonic screening; Challenges encountered at each of these stages, reasons for exclusion, and alternative recruitment strategies are presented and statistically described when possible. *Second*, we outline difficulties during the medical screening and the study visits, such as participants struggling to understand the task and complications in collecting blood/saliva biomarkers. *Third*, we discuss the importance of strong coordination and communication, both with participants and among study teams. Finally, specific guidelines are provided.

## 3 | Results and discussion: qualitative and descriptive statistics

This section summarizes key findings from the qualitative data and descriptive statistical analysis.

### 3.1. Recruitment and screening process

The participant recruitment and screening procedures are outlined below.

#### 3.1.1. | Eligibility criteria

Conducting a study including both a drug and a 7T magnetic resonance imaging (MRI), required careful consideration of several eligibility criteria. The most important was the diagnosis and characterization of the sample: we included participants diagnosed with MCI or mild dementia of the type Alzheimer’s dementia (AD). Diagnoses were made by medical doctors (co-authors of this paper, WG and MB) through a comprehensive clinical evaluation, including detailed anamnesis, neurological examination, cognitive testing, and cerebrospinal fluid (CSF) biomarkers, specifically, elevated tau pathology and reduced beta-amyloid levels. Individuals with mixed dementia were also eligible, provided that AD was a contributing diagnosis. All participants were required to be 60 years of age or older.

##### Eligibility criteria for Atomoxetine

For participants to be included, they had to be eligible for the study drug Atomoxetine/Strattera. Patients who take beta-blockers were excluded. Beta-blockers are a contraindication for Atomoxetine due to the possibility of experiencing cardiac arrhythmic events which may become fatal[5]. Additionally, all medications a patient was taking needed to be checked for interactions with Atomoxetine beforehand. Exclusion criteria also included a history of chemotherapy, brain radiation, a cancer diagnosis within the past two years or severe cardiac conditions and epilepsy or other forms of falling sickness, severe neurological and psychiatric diseases.

##### Eligibility criteria for 7T MRI

Participants had to fulfill strict 7T MRI eligibility criteria. Patients with implants (exception: dental implants) or metal in the body were not allowed to participate. In addition, patients who previously worked with metal or had any metal related accident were excluded [6]. Participants with tattoos were excluded because tattoo ink may contain metallic substances [7]. For each surgical procedure the participant had undergone, a doctor’s report was required. If the participant wore any other artificial objects such as artificial lenses, they had to provide documentation or a passport for the specific object. Additional exclusion criteria were claustrophobia or inability to lie still for an extended period of 2 hours. This ensured that the participants were not excessively stressed and that images did not contain movement artefacts.

#### 3.1.2. | Recruitment process

The recruitment process of the study was conducted at the memory clinic of the at the DZNE and the process was divided in two parts: The pre-selection and the telephonic screening. This elaborate process enabled us to maintain a sample that fit the study criteria well and ensured that only those patients who posed a minimal risk to any endangerment due to their health status were able to participate. Participants were pre-selected by studying doctor’s letters of patients from the memory clinic on site and excluding those that were ineligible. In the telephonic screening, the pre-selected participants were contacted and given an explanation of the study and, if the participant was interested, were asked questions following a questionnaire based on our eligibility criteria. If the participant passed the telephonic screening, they were invited to a medical screening in which specific data (i.e., cognitive / psychological tests, an electrocardiogram (ECG) and blood samples) were acquired to verify that the participants did not present depression or any blood or cardiological abnormality and to confirm that the memory was impaired. The medical screening was concluded by an examination performed by one of the doctors who confirmed if the patient was eligible for the study. If the patient passed the medical screening as well, we scheduled appointments for the study visits.

#### 3.1.3 | Descriptive stats of the recruitment process

Participants were recruited via our internal IKND database at the DZNE memory clinic. This included assessing the fit of patients to our study by systematically reviewing medical doctor’s letters and verifying compliance with the predefined inclusion criteria. As a disclaimer: By the time this article is being written, the recruitment process is ongoing. However, the recruitment data acquired so far is conclusive enough to present preliminary findings. In total, the memory clinic contains around 1000 active patients, from which 475 patients were part of the required demographic for our study (mild cognitive impairment or mild dementia of the Alzheimer’s dementia type). From this potential sample size, 44% (n = 211) had mild cognitive impairment, 55% had mild dementia (n = 263 and 1% (n = 1) had dementia but not a clear diagnosis yet. After pre-selection alone, we had to eliminate 49% (n = 235) of the patients due to various exclusion criteria they fulfilled (view Supplementary Table *1* for further details). So far, 44% (n = 211) of the patients from the demographic of interest have been contacted and 6% (n = 29) were not reachable or have not been contacted yet. Out of all patients contacted, 6% (n = 14) were a good fit for the MRI study, the rest either revealed certain exclusion criteria during the telephonic screening or were either not able to or not interested in participating. In regards of the total amount of patients fitting into our desired demographic, only 3% could be included in the study from the recruitment process.

As mentioned earlier, patients were excluded from the study due to various exclusion criteria they fulfilled. The most common reason was patients taking beta-blockers (30%), followed by patients not being interested in participating (20%), patients suffering from other health or mobility related problems which make study participation impossible (12%) and not being eligible for 7T MRI (7%). All reasons for exclusion and their distribution are summarized in Supplementary Table.

#### 3.1.4 | Challenges of recruitment

During recruitment, many challenges were faced which eventually led to the exclusion of many potential participants. Most patients were excluded due to admission of beta-blockers which is common in people of the age demographic of interest. Also, some patients already had implants or operations to which they did not have the doctor’s reports anymore, these participants were excluded as well. Having implants is a contraindication for participating in a 7T MRI scan[8]. Additionally, doctor’s reports for operations were a requirement for the MRI staff to safely exclude any possible implantation in the body. Furthermore, some patients could not be contacted because they did not answer their phones or had incorrect or invalid numbers in the database. Upon calling patients, they usually rejected participation in the study for multiple reasons. For instance, patients had other moderate to severe health conditions or impaired mobility which affected their possibilities to travel to the study site; some patients were not interested in taking part in a study. When conducting the telephonic screening with patients who did express an interest in the study, exclusion criteria were sometimes revealed during the telephonic screening and could not be extracted from studying the doctors’ letters alone; upon noticing further exclusion criteria, the telephonic screening had to be cancelled and the patient had to be excluded. These events led to a small number of participants that could potentially be included in the study. View *Table 1* in Supplementary Table.

#### 3.1.5 | Other ways of recruitment

Due to the high amount of exclusion criteria and the resulting difficulties of recruiting from the memory clinic alone, other ways of recruiting participants have been put to the test. For instance, other memory clinics nearby were contacted and asked for collaboration, however, none of them replied. Furthermore, the study was advertised by hanging posters and leaving flyers in doctors’ offices or clinics in the same city as the study site. That way, patients interested in the study could contact us themselves. Eventually only internal patients were included. Although limiting recruitment to the local memory clinic made the process slower due to fewer available participants, it came with important advantages. It ensured consistent diagnoses across all participants and guaranteed that we had Alzheimer’s disease biomarkers (CSF Tau and β-amyloid) for all our included participants. This led to a well-characterized and reliable study sample.

### 3.2. | Challenges during the study

If the participants who successfully passed the recruitment process were invited to a medical screening in which initial tests were administered, and ECG data and blood samples were collected. The administered tests include the *Montreal Cognitive Assessment* [9] to assess the severity of cognitive impairment and the *Geriatric Depression Scale* to assess identify depressive symptoms [10]. The medical screening was concluded by an examination by one of the doctors who finally assessed if the patient was able to participate in the study. If the patient passed the medical screening as well, they were given appointments for the study visits.

#### 3.2.1 | Challenges during the medical screening

Medical screenings after successful recruitment still presented some challenges. For instance, many participants were talkative, which was problematic due to the limited time during the screening. Another time-consuming factor was taking blood. This was especially challenging with participants who had small or hardly visible veins. Additionally, some participants had trouble understanding the study. As dementia can present itself in different ways and stages, it is to be expected that some participants, depending on the manifestation of their illness, are not able to fully understand the goal and means of the study. This was the case when participants, for instance, were not able to recall what the study was about after they were told about the goal and procedures of the study or when participants repeatedly gave inadequate answers to questions. If during the screening a participant was not able to fully understand the subject of the study or was unfit in other ways, the assumption was made that the participant was not able to give informed consent and was therefore excluded from the study.

Sometimes, arguments for exclusion of the participant were not revealed until the medical screening. An example would be the participant remembering a previous operation during the screening without a report being available. To not waste time and resources researchers are strongly advised to screen the participant in the recruiting process thoroughly and ask about different kinds of operations specifically. For instance, instead of asking if the participant has had any operations in general, try asking about as many specific operations as possible (such as eye-operation, colon-operation, nose-operation, operation due to fracture, etc.). People with memory impairments might not remember procedures they had if asked generally, conversely, specific questions provide the participant with a keyword that makes it easier to recall a procedure they possibly went under.

#### 3.2.2. | Challenges during the study visits

If the recruiting and screening process is not done thoroughly, specific exclusion criteria might be revealed as late as during the first study visit. This happened with some participants who did not disclose previous procedures during the telephonic and medical screening and remembered them during the first visit. This case is best to be avoided because cancelling MRI appointments last-minute is usually expensive. Therefore, it is imperative that exclusion criteria is checked carefully and, if needed, multiple times.

Some participants experienced side effects due to the MRI. Side effects might include a rising temperature of tissue in the MRI, headaches, dizziness or a tingling sensation in certain body parts[11]. Each person reacts differently to possible side effects, a few participants however experienced strong discomfort during an MRI scan and wished to discontinue the study.

While a participant was lying in the MRI scanner, they were assigned a task for which they had to react to stimuli presented on a screen inside the scanner by pressing buttons on remotes in their hands in accordance with the presented stimuli. The task had to be explained by a research assistant via intercom system between the control room and the MRI scanner. This was challenging due to the noise of the MRI scanner, especially if participants were hard of hearing, therefore the task had to be explained slowly and with appropriate emphasis, using simple sentences and as few words as possible. A possible way to avoid this challenge is by explaining the instructions before participants enter the scanner. This does not apply to people with more severe memory impairments, however, because previously explained instructions are usually forgotten by the time the participant enters the scanner. It is advisable that the task in the scanner contains a written instruction as well so that the participant can read the instruction before performing the task. Because it is hard for some people to read the instructions due to the unusual circumstances, even with MRI-safe reading glasses provided by the staff, it is best that a staff member explains the instructions in addition.

One of the tasks included in the fMRI paradigm is the well-known Stop Signal Task (SST). Originally introduced by Lappin and Eriksen in 1966 [12] and further developed by Logan and Cowan in 1984 [13], the SST has been widely used to investigate reaction times and executive functions, particularly inhibitory control. In this task, participants are presented with various stimuli to which they must respond, inhibit their response, or withdraw an already initiated response (see Figure 1 and Supplementary Material).

**Figure 1.**
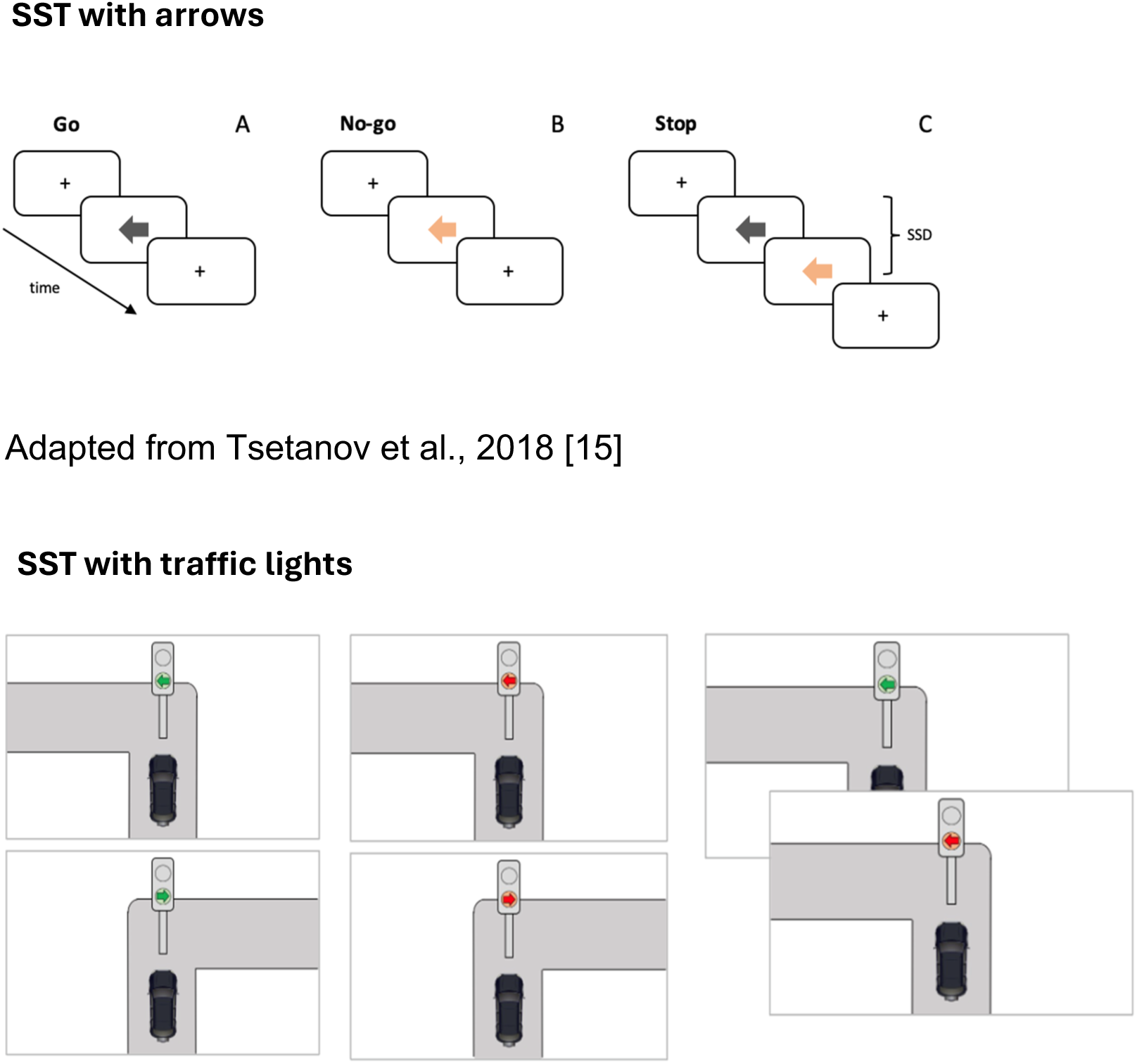
Stop signal task stimuli presentation. Note. Figure 1 shows two different stimuli type of a stop signal task (SST): the SST with arrows is adequate for young adults and the SST with traffic lights for older adults and especially older adults with mild cognitive impairment (MCI).

The specific nature of the stimuli is especially important for participants with Mild Cognitive Impairment (MCI) or Alzheimer’s Disease (AD), as it should be as intuitive and visual as possible to ensure task comprehension. Different type of SST stimuli were previously assessed in different lab studies, concluding that the type of stimuli and “story” behind them can facilitate the understanding of it, e.g., presenting the task as a scene were the participant is driving, sees a traffic light and has to decide to continue driving (press mouse button), stop (do not press mouse button) or withdraw the initial decision of “walking” [14] because the light turned red in contrast to simply presenting “decontextualized arrows” makes a big difference and helps considerably in understanding the task (Figure 1).

Additionally, the SST is designed so that participants succeed on approximately 50% of trials. If participants adopt strategies, such as deliberately delaying responses to increase their chances of success, this can alter the neural mechanisms being engaged, shifting the task from one of inhibitory control to one of decision-making. It is relevance to include “a practice SST” before the scan session in the study protocol. This way it can be ensured the participant has understood well the instructions.

Therefore, it is crucial to provide clear and appropriate instructions, emphasizing that the goal is not to perform “perfectly.” This is particularly important for older adults and those with MCI, who may feel compelled to compensate for their cognitive difficulties by trying to achieve overly high performance (see Supplementary Material). Another task included in the study was the incidental emotional memory task, in which participants viewed a series of scenes during the fMRI scan and, one day later, completed a recognition memory task, outside the scan. The task was well-suited to our study aims; however, participants with more advanced cognitive impairment or mild dementia occasionally experienced difficulty responding within the given time, even when they recognized the images. To address this, a research assistant was available to provide support, when necessary, e.g., the participant verbally responded and the assistant pressed the mouse button for them. This approach ensured that the task continued to primarily assess memory performance rather than processing speed, in line with the study’s objectives.

### 3.3. | Relevance of coordination and communication with the participants and across the study teams

Conducting the study required communication with different parties, especially when scheduling appointments for visits or medical screenings. Prior to contacting patients, the availabilities of research assistants who were conducting the appointments as well as doctors who were assessing the participants in the medical screening had to be checked. For the study visits, the appointments were given by the MRI technicians and at least two research members were made responsible for conducting the visits. The appointments were made in accordance with the schedules of the research team, the doctors, the participants and, if necessary, their caregivers and scheduled appointments had to be reported to the receptionist of the memory clinic who kept an overview of the doctors’ schedules and availabilities.

In some cases, communication needed to involve not only the patients but also their caregivers. This was especially important when patients had trouble remembering appointments or medical history, or when a caregiver’s input was required for one of the questionnaires. Caregivers also played a vital role in helping patients with mobility issues get to the study site.

Clear and transparent communication within the study team was crucial due to the high number of patients that needed to be pre-selected, contacted, and screened. A shared patient table was maintained and regularly updated with contact details, diagnosis, exclusion criteria, and current status—such as agreement to participate, no response, requests to reschedule, or refusals. This allowed all team members to stay informed and coordinate efficiently.

While team members had defined roles, such as recruitment or preparing study materials, everyone needed to stay updated on participant status, recruitment progress, upcoming tasks, and any challenges. This ensured smooth collaboration, minimized errors, and supported continuous improvement.

### 3.4. | Conclusions and guidelines for researchers

As a conclusion, conducting a pharmacological 7T MRI study with Alzheimer’s population poses many challenges which can require adaptation of methods or eligibility criteria. Compared to a 7T MRI, a 3T MRI reduces the safety concerns and subjective complaints[16]. The lower magnetic field of the 1.5 and 3T MRI is usually associated with reduced adverse effects, such as vertigo, dizziness of participants[16,17] and sensation of heating or metallic taste. If a 3T or 1.5T MRI is used instead of a 7T MRI, eligibility criteria might be less conservative, allowing for broader participant inclusion. However, this comes at the cost of reduced spatial resolution and sensitivity to capture small structures. Given that no serious adverse events have been reported at either field strength, and that the ultra-high field MRI is very well tolerated, only around a 3% of individuals report discomfort in 7T MRI [11,18], the decision to use 3T or 7T should be based on the specific aims of the study and resources available, i.e., time, economic resources and infrastructure[19]. A pharmacological intervention may also increase the difficulty of conducting a study and recruit eligible participants due to adverse interactions of other medications. In the case of this study, atomoxetine had an adverse interaction with beta-blockers which patients take due to high blood pressure or other cardiological diseases. Although beta-blockers generally do not cross the blood-brain barrier, atomoxetine may counteract their effects, potentially putting the participant at risk. Since cardiovascular diseases are common in the age group relevant to this study, a large number of potential participants had to be excluded. Administering a study drug with little side effects or less relevant adverse interactions with other medications might facilitate the recruitment process.

Based on the considerations outlined throughout this article so far, we present a set of guidelines for conducting clinical MRI studies involving pharmacological interventions in populations with Alzheimer’s population. These recommendations are intended to give an orientation to researchers designing studies with similar methods or populations.

- **Thoroughly screen each participant for relevant exclusion criteria for the MRI before inviting them to a study visit. If unsure, double-check by asking the patient more questions or, if need be, ask the doctors or caregiver responsible for the patient.**

Following this guideline avoids additional costs due to recruitment of participants who fulfill exclusion criteria that were overlooked by not screening the participant carefully beforehand.

- **Communicate clearly with each party involved in the study: Participants, doctors, caregivers, MRI technicians as well as researchers within the study team itself.**

By doing so, uncertainties, missing information and misunderstandings can be avoided.

- **Explain study relevant information to the participants in a way they can understand.**

Individuals with Alzheimer’s disease experience memory impairment and depending on their educational background, might have a hard time understanding medical or technical terms. Try explaining the aim and process of the study while avoiding overly medical or technical terms which require an understanding that exceeds basic education. If such terms are used, make sure that the participant understands what they mean by asking if they are familiar with the term and give an explanation if needed.

- **Be patient and understanding with participants but also remember to remain professional.**

Due to memory impairments in Alzheimer’s dementia, participants sometimes need to be reminded of appointments or task instructions. Be ready to provide those multiple times if need be. Most patients who participated were more than happy to give information we asked for, which sometimes resulted in the participant talking for an extended period. While it is advised to keep an open ear for talkative patients, it is also important to keep the time you have in mind, especially if it is limited. If the participant strays from the subject, try guiding them back on topic gently and politely. Being aware of dementia as an incurable disease, some patients struggle with accepting it as part of themselves or are emotionally distressed due to it. These participants also might have the need to express their experiences and if they do, do not cut them off rapidly but instead, try to listen, show understanding but remain on the subject by, for instance, reinforcing their participation in our study.

- **Try to not get discouraged by stagnation in the recruitment process and aim towards expanding recruiting strategies, if necessary.**

The recruitment process for a medical 7T MRI study with Alzheimer’s population involving a study drug is challenging due to the number of eligibility criteria that needs to be considered (see Supplementary Table), a large percentage of possible participants had to be excluded from the study due to various reasons. Sometimes, this can call for expansion of recruitment strategies. If recruiting participants from your source of choice becomes increasingly difficult, try utilizing other means such as cooperating with other medical facilities to get access to more possible participants.

- **Use tasks and stimuli that are appropriate for the cognitive level of your study population.**

Some fMRI paradigm tasks were originally developed for basic neuroscience research involving younger and/or healthy participants. Therefore, careful selection, piloting, and adaptation of these tasks to the specific study population are essential to ensure that participants fully understand the instructions and that the task accurately measures the intended cognitive function.

- **Train research assistant staff accordingly and carefully.**

If you are the study director and you hire staff as research assistants, make sure you carefully train them in each aspect of their respected assignments. Familiarize staff with study protocols, study information and consent form, all eligibility criteria, selection, screening and recruitment processes. Emphasize the importance of transparent communication with all parties involved, make sure to regularly review the progress together and reflect on possibilities to enhance the workflow, if necessary.

## Data Availability

All data produced in the present study are available upon reasonable request to the authors

## Acknowledgments

The authors would like to gratefully acknowledge all the participants who kindly provided their time and effort.

## Sources of Funding

This work was supported by Sonderforschungsbereich 1315, Project B06, The Alzheimer’s Research United Kingdom (ARUK) SRF2018B-004, NIH R01MH126971.

## Consent statement

Informed consent was obtained from all subjects involved in this study.

## Data Availability Statement

Further details on the data are available upon request. Please contact Lucía Penalba-Sánchez at the Institute of Cognitive Neurology and Dementia Research (IKND), German Center for Neurodegenerative Diseases (DZNE), Magdeburg.

## Disclosures

Conflict of interest statement: all authors certify that they have no affiliations with or involvement in any organization or entity with any financial or non-financial interest in the subject matter or materials discussed in this manuscript.

## Notes

### Competing Interest Statement

The authors have declared no competing interest.

### Funding Statement

This work was supported by Sonderforschungsbereich 1315, Project B06, The Alzheimers Research United Kingdom (ARUK) SRF2018B-004, NIH R01MH126971.

### Author Declarations

This study was approved by the local Ethics Committee of the Otto-von-Guericke University Magdeburg (reference number 190/19). All participants provided written informed consent prior to participation, in accordance with the Declaration of Helsinki.

