## Supplementary material for "Conducting MRI trials in Alzheimer’s patients: Challenges and Guidelines": Supplementary Material.docx

**Information on our case study:**

Our study aimed to investigate the effects of atomoxetine on cognitive performance and memory retention by performing tests with participants, additionally conducting a brain scan with magnetic resonance imaging (MRI) and implementing a control trial with placebo. Atomoxetine or Strattera is medication commonly used to treat attention-deficit-/hyperactive disorder (ADHD), however, studies suggest that this drug can also improve memory and cognitive performance in people with memory impairments. For further information of the study do not hesitate to contact the correspondence author Lucía Penalba-Sánchez.

**SST Instructions:**

The instructions below were read to the participants before performing the Stop Signal Task (SST). After explaining the instructions, participants are asked to explain the task back to us to ensure they understood it properly.

"In this task, imagine you are driving a car. When you see a green traffic light pointing to the right, press the right mouse (any right button). If you see a green traffic light pointing to the left, press the left mouse (any left button).” ***Note for assistants:*** *Show the participant the mouse(s) in the scan to help them understand that each mouse (right/left) has two buttons, and it does not matter which one they press.*

“When you see a red traffic light, do not press any button. Sometimes, you will see a green traffic light that suddenly changes to red; in this case, do not press any button. In this task, aim for both accuracy and speed. While accuracy is important, we also value speed. It's okay to make mistakes; being 100% right is not the objective. If you make some mistakes, you are doing well! Never wait too long; otherwise, we may not be able to use your data. Being right does not mean being 100% right, but being half accurate and fast! We penalize perfection!

***Note for assistants:*** *Between blocks remember to the participant that the goal is not being perfect and that it is important to also be fast, so making some mistakes it is a very good sign!). After the practice part and after the first block we check the data to see how it is going and we give feedback / instructions again depending on how they have been doing during the first block.*

Figure 1. Breakdown of reasons for excluding patients from the study:

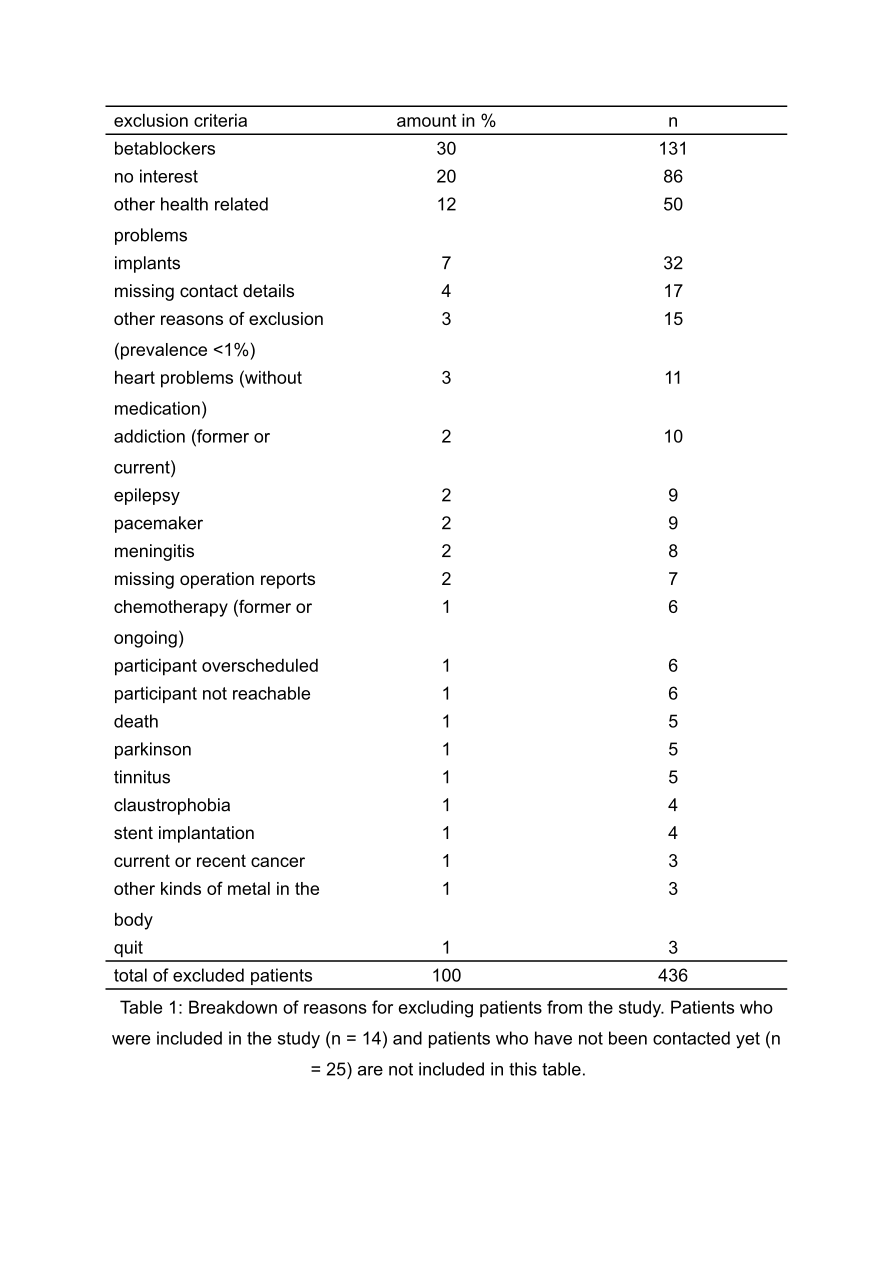
